# A New Predictor of Disease Severity in Patients with COVID-19 in Wuhan, China

**DOI:** 10.1101/2020.03.24.20042119

**Authors:** Ying Zhou, Zhen Yang, Yanan Guo, Shuang Geng, Shan Gao, Shenglan Ye, Yi Hu, Yafei Wang

## Abstract

**Background:** Severe acute respiratory syndrome coronavirus 2 (SARS-CoV-2) broke out in Wuhan, Hubei, China. This study sought to elucidate a novel predictor of disease severity in patients with coronavirus disease-19 (COVID-19) cased by SARS-CoV-2.

**Methods:** Patients enrolled in this study were all hospitalized with COVID-19 in the Central Hospital of Wuhan, China. Clinical features, chronic comorbidities, demographic data, and laboratory and radiological data were reviewed. The outcomes of patients with severe pneumonia and those with non-severe pneumonia were compared using the Statistical Package for the Social Sciences (IBM Corp., Armonk, NY, USA) to explore clinical characteristics and risk factors. The receiver operating characteristic curve was used to screen optimal predictors from the risk factors and the predictive power was verified by internal validation.

**Results:** A total of 377 patients diagnosed with COVID-19 were enrolled in this study, including 117 with severe pneumonia and 260 with non-severe pneumonia. The independent risk factors for severe pneumonia were age [odds ratio (OR): 1.059, 95% confidence interval (CI): 1.036–1.082; p < 0.001], N/L (OR: 1.322, 95% CI: 1.180–1.481; p < 0.001), CRP (OR: 1.231, 95% CI: 1.129–1.341; p = 0.002), and D-dimer (OR: 1.059, 95% CI: 1.013–1.107; p = 0.011). We identified a product of N/L*CRP*D-dimer as having an important predictive value for the severity of COVID-19. The cutoff value was 5.32. The negative predictive value of less than 5.32 for the N/L*CRP*D-dimer was 93.75%, while the positive predictive value was 46.03% in the test sets. The sensitivity and specificity were 89.47% and 67.42%. In the training sets, the negative and positive predictive values were 93.80% and 41.32%, respectively, with a specificity of 70.76% and a sensitivity of 89.87%.

**Conclusions:** A product of N/L*CRP*D-dimer may be an important predictor of disease severity in patients with COVID-19.

## Introduction

Since last December, a growing cluster of pneumonia cases caused by the 2019 novel coronavirus was reported in Wuhan, Hubei Province, China. The virus rapidly spread from Wuhan to the surrounding provinces and cities, which attracted worldwide attention and public panic^1-3^. Eventually, the pneumonia was designated as coronavirus disease-19 (COVID-19) by the World Health Organization (WHO) on February 11, 2020. The International Committee on Taxonomy of Viruses declared that the virus was officially classifiable as severe acute respiratory syndrome (SARS) coronavirus 2 (SARS-CoV-2) on the same day. SARS-CoV-2 belongs to the genus beta coronavirus of the *Coronaviridae* family, which includes the deadly SARS and Middle East respiratory syndrome coronaviruses^4-6^. Studies have shown that SARS-CoV-2 displays roughly an 89% homology with bat SARS-like-CoVZXC21 and 82% homology with human SARS-CoV^7^. It is highly contagious and has a high mortality rate^8^. As of 22:00 on March 10, 2020, there were 80,933 confirmed cases, 4,794 severe cases, and 3,140 deaths in China as well as 35,606 confirmed cases and 975 deaths collectively among all other countries.

The disease is prone to causing respiratory symptoms, such as high fever, dyspnea, and cough. It may rapidly develop into severe pneumonia, acute respiratory distress syndrome (ARDS), multiple organ dysfunction syndrome, and death^9^. As the number of patients with COVID-19 increases, so too does the number of severe cases. Thus far, there are no drugs approved or verified that are specific to the virus^10, 11^. Thus, the treatment of patients critically ill with COVID-19 is becoming one of the main challenges facing clinicians and there is a need adopt reliable predictors of the severity of COVID-19 to identify and treat the most severe patients in the early stage. This study therefore sought to explore the adoption of a factor for the prediction of severity in patients with COVID-19 and provide evidence of its use for the screening of severely ill patients.

## Methods

### Patients

Patients enrolled in this study were hospitalized at the Central Hospital of Wuhan, which is a tertiary teaching hospital and which is responsible for the treatment for patients of COVID-19, as assigned by the Chinese government. All of the patients were local residents of Wuhan and had contact with people who had been confirmed (or suspected) to have contracted the illness. The relevant laboratory examinations, common pathogen detection, and chest computed tomography (CT) scans were completed. Each patient also underwent the SARS-CoV-2 nucleic acid swab test, with some patients testing positive and others testing negative. The immaturity of the methods and an incorrect sampling method used for SARS-CoV-2 nucleic acid detection may have contributed to the false-negative results. High-resolution CT scans with a scan layer thickness of 5 mm and a reconstruction of a 1- to 1.5-mm thick layer are recommended for the radiological examination of COVID-19. The included patients were clinically diagnosed with COVID-19 according to the WHO’s interim guidance^12^. For patients who were suspected to have the illness, two senior respiratory doctors made the diagnosis together.

The study participants were divided into two groups: patients with severe pneumonia and those with non-severe pneumonia. Patients with the following clinical signs were considered to have severe pneumonia: fever or suspected respiratory infection plus either a respiratory rate of greater than 30 breaths/min, severe respiratory distress, or SpO_2_ of less than 90% on room air. Patients with ARDS, sepsis, or septic shock were also included. Those patients without the above severe signs were included in the non-severe pneumonia group. Patients with other viral, bacterial, and fungal infections upon admission and those with missing data were excluded. This study was approved by the ethics committee of Wuhan Central Hospital. As the study is a retrospective study and does not involve patients’ privacy, the informed consent can be exempted

### Data collection

Chronic comorbidities, demographic data, laboratory examinations, and chest CT scans were reviewed using electronic medical records. Data on white blood cell (WBC) count and lymphocyte (L), neutrophil (N), alanine transaminase (ALT), aspartate aminotransferase (AST), serum albumin (ALB), serum creatinine (Cr), blood urea nitrogen (BUN), D-dimer, and C-reactive protein (CRP) levels were included. These data were acquired by physicians and were the results of an examination on the first day after admission. All of the data were checked by another researcher to ascertain their accuracy.

### Internal validation

Patients were randomly assigned in a 2:1 fashion to undergo training and hold-out test sets. The predictors were trained in the training group and the optimal cutoff value was obtained.

This internal validation assessed the value for predicting the severity of with COVID-19. Statistical performance for the classifier was measured by calculating sensitivity, specificity, and negative and positive predictive values.

### Statistical analysis

The continuous variables were either presented as means and compared using t-tests if they were normally distributed or were described using medians. The Mann–Whitney U test was used for comparisons. Categorical variables were presented as count (%) and compared by the chi-squared test or Fisher’s exact test. Logistic regression analysis was used to assess risk factors of severe pneumonia. A receiver operating characteristic (ROC) curve was used to filter predictors, and internal validation was used to validate the predictive value of predictors. A two-sided α value of less than 0.05 was considered to be statistically significant. We used the Statistical Package for the Social Sciences, version 22.0 (IBM Corp., Armonk, NY, USA) for statistical analysis.

## Results

### Baseline characteristics

Between January 1 and February 28, 2020, a total of 393 patients with COVID-19 were enrolled, but only 377 eligible participants were finally included in our analysis. Among them, there were 172 (45.62%) patients with positive SARS-CoV-2 nucleic acid test results, 106 (40.93%) patients with non-severe pneumonia, and 66 (56.41%) patients with the severity of COVID-19. The basic characteristics of the study population are shown in Table 1. The patients with severe pneumonia tended to older. Also, patients with chronic comorbidities, such as high blood pressure (HBP), type 2 diabetes mellitus (T2DM), coronary heart disease (CHD), and chronic obstructive pulmonary disease, accounted for a larger proportion of the population in the severe pneumonia group. Relative to those with non-severe pneumonia, patients with severe pneumonia had significantly high serum WBC count and N, CRP, Cr, BUN, D-dimer levels and lower albumin and lymphocyte levels. The ratio of neutrophils to lymphocytes (N/L) was significantly higher in patients with severe pneumonia. In addition, no significant differences in ALT, AST, or time between onset and admission were apparent.

**Table 1:**
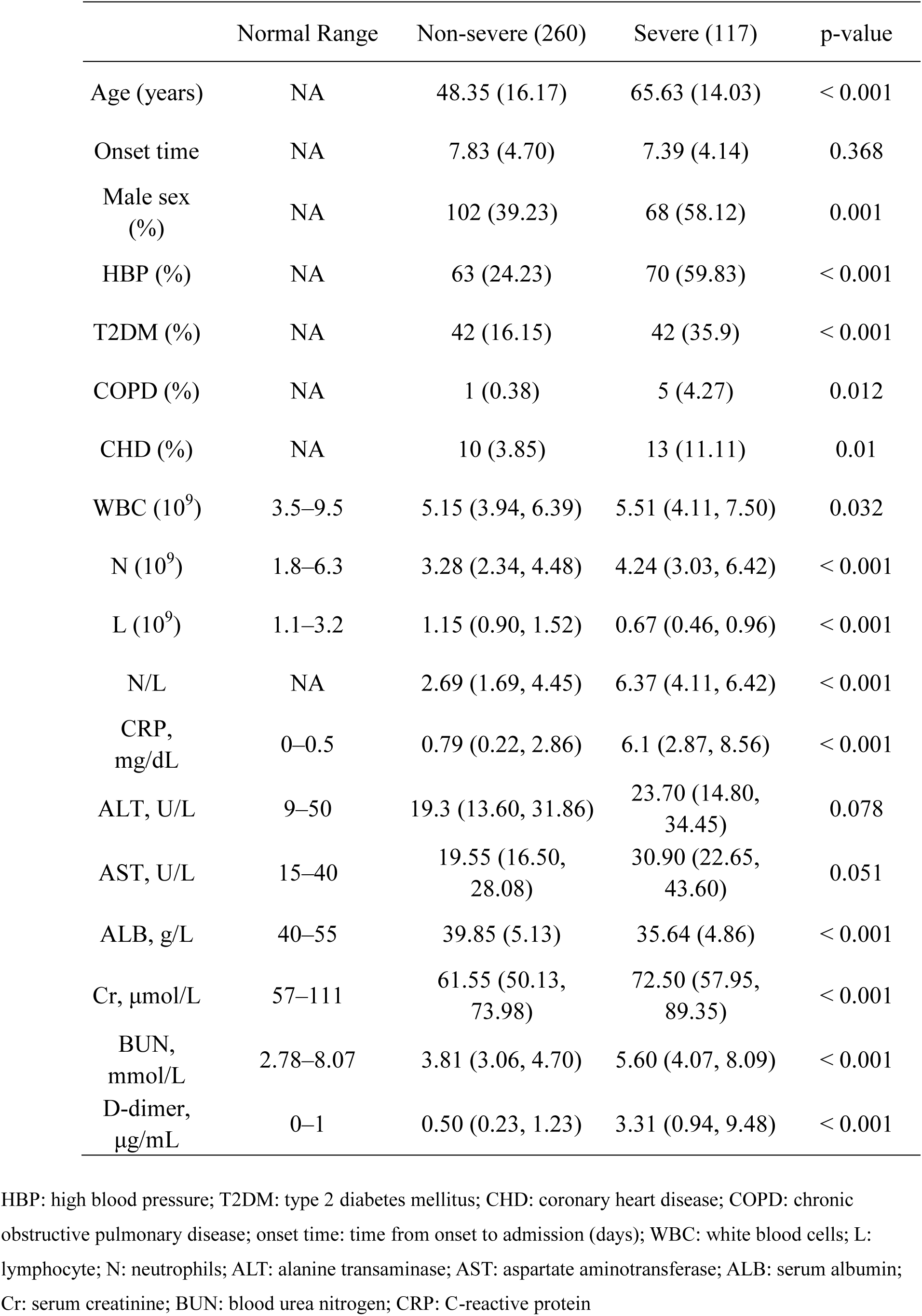
Baseline characteristics of the study participants

|  | Normal Range | Non-severe (260) | Severe (117) | p-value |
| --- | --- | --- | --- | --- |
| Age (years) | NA | 48.35 (16.17) | 65.63 (14.03) | < 0.001 |
| Onset time | NA | 7.83 (4.70) | 7.39 (4.14) | 0.368 |
| Male sex (%) | NA | 102 (39.23) | 68 (58.12) | 0.001 |
| HBP (%) | NA | 63 (24.23) | 70 (59.83) | < 0.001 |
| T2DM (%) | NA | 42 (16.15) | 42 (35.9) | < 0.001 |
| COPD (%) | NA | 1 (0.38) | 5 (4.27) | 0.012 |
| CHD (%) | NA | 10 (3.85) | 13 (11.11) | 0.01 |
| WBC (10<sup>9</sup>) | 3.5–9.5 | 5.15 (3.94, 6.39) | 5.51 (4.11, 7.50) | 0.032 |
| N (10<sup>9</sup>) | 1.8–6.3 | 3.28 (2.34, 4.48) | 4.24 (3.03, 6.42) | < 0.001 |
| L (10<sup>9</sup>) | 1.1–3.2 | 1.15 (0.90, 1.52) | 0.67 (0.46, 0.96) | < 0.001 |
| N/L | NA | 2.69 (1.69, 4.45) | 6.37 (4.11, 6.42) | < 0.001 |
| CRP, mg/dL | 0–0.5 | 0.79 (0.22, 2.86) | 6.1 (2.87, 8.56) | < 0.001 |
| ALT, U/L | 9–50 | 19.3 (13.60, 31.86) | 23.70 (14.80, 34.45) | 0.078 |
| AST, U/L | 15–40 | 19.55 (16.50, 28.08) | 30.90 (22.65, 43.60) | 0.051 |
| ALB, g/L | 40–55 | 39.85 (5.13) | 35.64 (4.86) | < 0.001 |
| Cr, µmol/L | 57–111 | 61.55 (50.13, 73.98) | 72.50 (57.95, 89.35) | < 0.001 |
| BUN, mmol/L | 2.78–8.07 | 3.81 (3.06, 4.70) | 5.60 (4.07, 8.09) | < 0.001 |
| D-dimer, µg/mL | 0–1 | 0.50 (0.23, 1.23) | 3.31 (0.94, 9.48) | < 0.001 |

HBP: high blood pressure; T2DM: type 2 diabetes mellitus; CHD: coronary heart disease; COPD: chronic obstructive pulmonary disease; onset time: time from onset to admission (days); WBC: white blood cells; L: lymphocyte; N: neutrophils; ALT: alanine transaminase; AST: aspartate aminotransferase; ALB: serum albumin; Cr: serum creatinine; BUN: blood urea nitrogen; CRP: C-reactive protein

### Risk factors of severe pneumonia

To further determine the independent risk factors of severe pneumonia, we performed a logistic regression analysis. The results indicated that, after adjusting for confounding factors such as gender, T2DM, HBP, CHD, Cr, BUN, and ALB, the independent risk factors for severe pneumonia were age [odds ratio (OR): 1.059, 95% confidence interval (CI): 1.036–1.082; p < 0.001], N/L (OR: 1.322, 95% CI: 1.180–1.481; p < 0.001), CRP (OR: 1.231, 95% CI: 1.129–1.341; p = 0.002), and D-dimer (OR: 1.059, 95% CI: 1.013–1.107l p = 0.001) (Table 2).

**Table 2.** The logistic regression of risk factors for the severity of COVID-19

|  | OR | 90% CI | p-value |
| --- | --- | --- | --- |
| Age | 1.059 | (1.036–1.082) | < 0.001 |
| N/L | 1.322 | (1.180–1.481) | < 0.001 |
| CRP | 1.231 | (1.129–1.341) | 0.002 |
| D-dimer | 1.059 | (1.013–1.107) | 0.011 |

HBP: high blood pressure; T2DM: type 2 diabetes mellitus; CHD: coronary heart disease; L: lymphocyte; N: neutrophils; ALB: serum albumin; Cr: serum creatinine; BUN: blood urea nitrogen; CRP: C-reactive protein. N/L: the ratio of neutrophils to lymphocytes;
The logistic regression model included the following factors: age, gender, T2DM, HBP, CHD, Cr, BUN, N/L, CRP, D-dimer, and ALB.

### Predictors of severity pneumonia

The ROC curve was used to analyze the predictive value of N/L, CRP, and D-dimer for determining disease severity in patients with COVID-19. The area under the ROC curve (AUC) was further estimated (Figure 1-3), which suggested that these factors were not ideal predictors of the severity of COVID-19. To find a better predictor, we combined the three variables in different ways and analyzed the ROC curve for the new factors. Finally, the result showed that the product of N/L, CRP, and D-dimer (N/L*CRP*D-dimer) was a factor worth exploring. We performed an ROC curve analysis of N/L*CRP*D-dimer for diagnosing severe pneumonia. The AUC was 0.888 (95% CI: 0.856–0.921), with a specificity of 65.3% and a sensitivity of 95.7% (Figure 4). The cutoff value was 2.6839.

**Figure 1.**
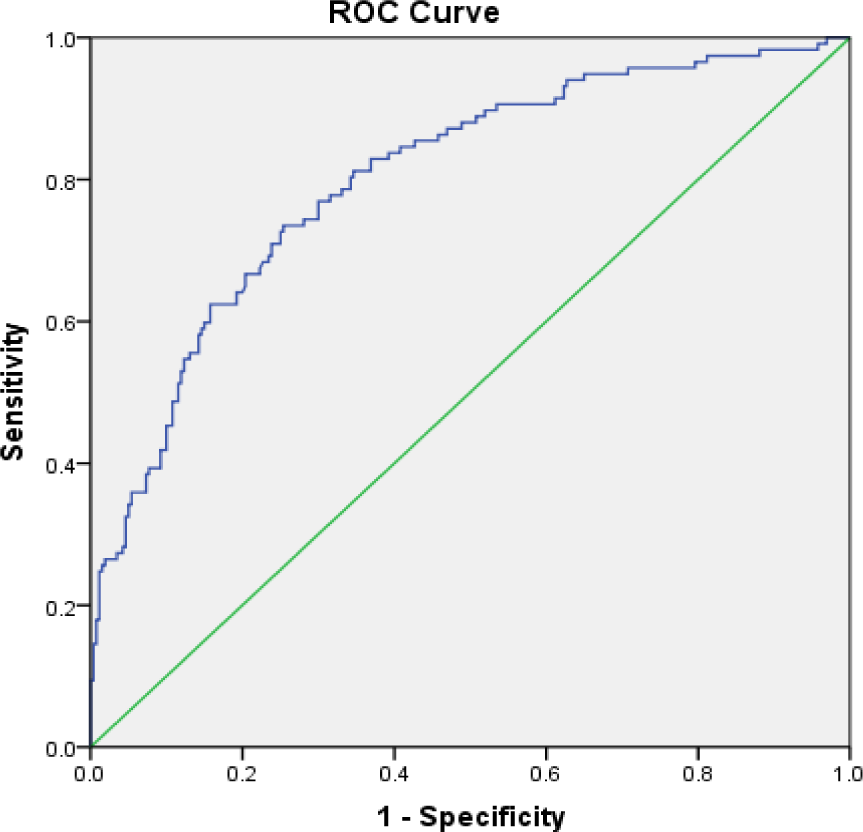
The receiver operating characteristic curve for N/L diagnosis of the severity of COVID-19; AUC: 0.802 (95% CI: 0.753–0.850).

**Figure 2.**
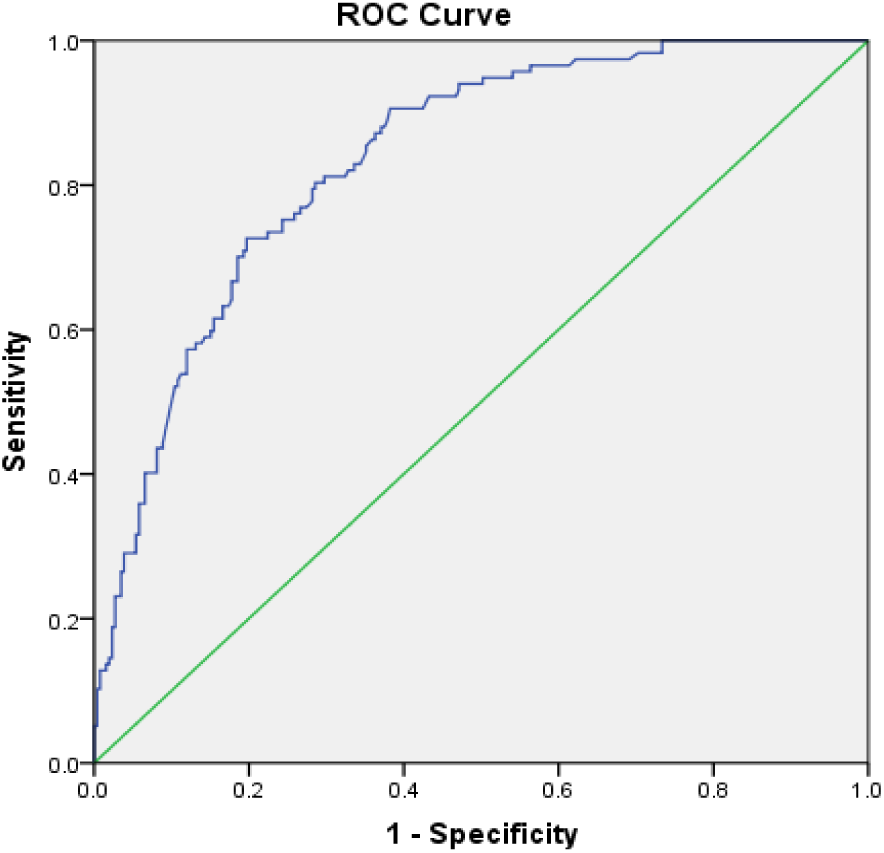
The receiver operating characteristic curve for CRP diagnosis of the severity of COVID-19; AUC: 0.836 (95% CI: 0.795–0.877).

**Figure 3.**
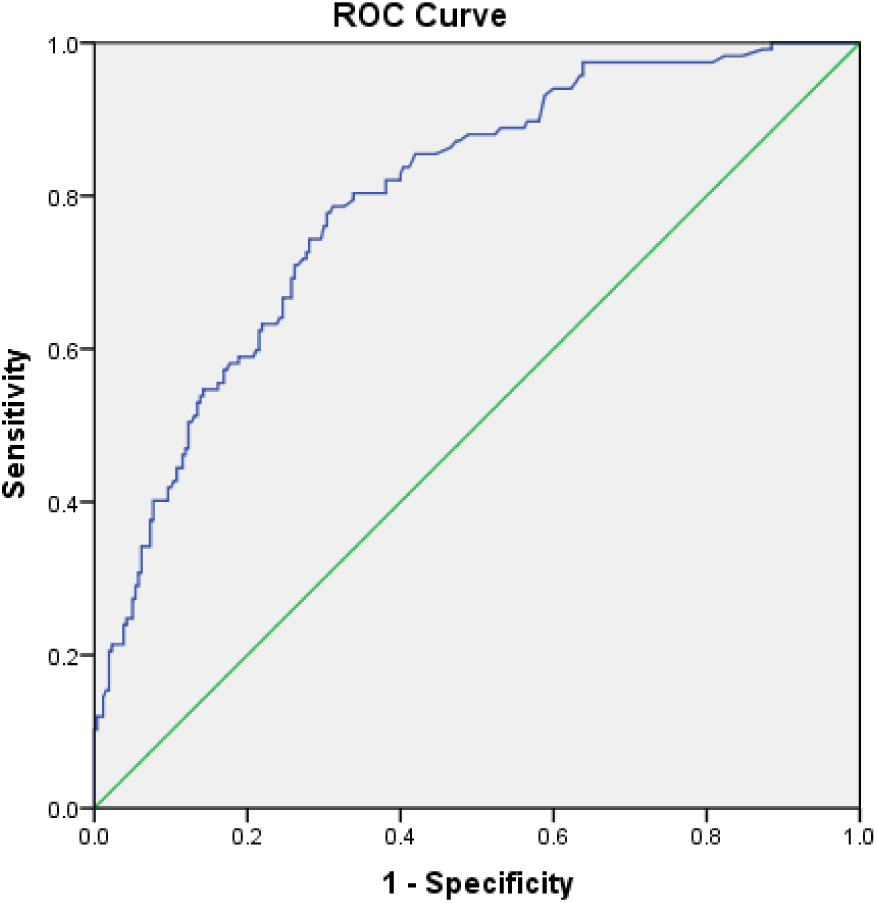
The receiver operating characteristic curve for D-dimer diagnosis of the severity of COVID-19; AUC: 0.795 (95% CI: 0.748–0.842).

**Figure 4.**
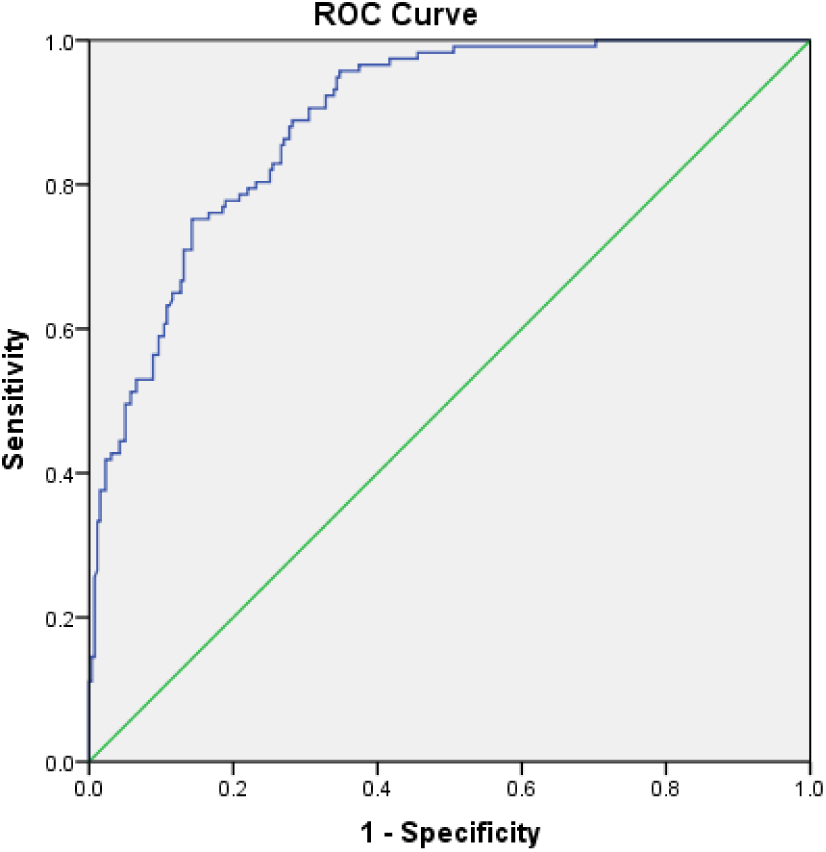
The receiver operating characteristic curve for N/L*CRP*D-dimer diagnosis of the severity of COVID-19; AUC: 0.888 (95% CI: 0.856–0.921).

### Internal validation

To further explore the predictive value of N/L*CRP*D-dimer in the severity of COVID-19, we also conducted an internal validation. About 30% of patients were randomly selected from the total population as test sets and the rest were designated as the training sets. Basic characteristics of patients in the two sets are listed in Table 3. ROC curves for the new predictor in the test sets are shown in Figure 5; the cutoff point was not derived. The AUC was 0.879 (95% CI: 0.838–0.921), with a specificity of 73.7% and a sensitivity of 88.6% in the training sets (Figure 6), and the cutoff value was 7.9622. The optimal cutoff value obtained from the average cutoff value of the original data group and the training sets was 5.32; thus, patients in the training and test sets were reclassified according to this result. Patients with N/L*CRP*D-dimer results of less than 5.32 were classified as patients with non-severe pneumonia in predicted groups. Otherwise, they were classified as patients with severe pneumonia. As can be seen from Tables 4 and 5, the negative predictive value (a diagnosis of non-severe pneumonia) of less than 5.32 for the N/L*CRP*D-dimer was 93.75% (95% CI: 0.8456–0.9799), and the positive predictive value (a diagnosis of severe pneumonia) was 46.03% (95% CI: 0.3431–0.5821) among the test sets. The sensitivity and specificity were 89.47% (95% CI: 0.7529–0.9641) and 67.42% (95% CI: 0.5711–0.7628), respectively.

**Table 3:** Baseline characteristics of patients in the training and test sets

|  | Training sets |  |  | Test sets |  |  |
| --- | --- | --- | --- | --- | --- | --- |
|  | Non-severe<br>(171) | Severe (79) | p-value | Non-severe (89) | Severe (38) | p-value |
| Age (years) | 49.05 (16.61) | 65.23 (15.00) | < 0.001 | 47 (15.29) | 66.47 (11.90) | < 0.001 |
| Onset time | 7.61 (4.72) | 7.15 (3.83) | 0.447 | 8.24 (4.79) | 7.89 (4.75) | 0.707 |
| Male sex<br>(%) | 66 (38.6) | 45 (56.96) | 0.009 | 36 (40.45) | 23 (60.53) | 0.052 |
| HBP (%) | 40 (23.39) | 49 (62.03) | < 0.001 | 23 (25.84) | 21 (55.26) | 0.002 |
| T2DM (%) | 29 (17) | 29 (36.71) | 0.001 | 13 (14.61) | 13 (34.21) | 0.017 |
| CHD (%) | 7 (4.09) | 5 (6.33) | 0.145 | 3 (3.37) | 6 (15.79) | 0.021 |
| WBC (10<sup>9</sup>) | 5.13 (3.78, 6.42) | 5.51 (4.00, 7.30) | 0.118 | 5.20 (3.99, 6.30) | 3.43 (4.47, 7.76) | 0.142 |
| N (10<sup>9</sup>) | 3.25 (2.32, 4.55) | 4.24 (3.01, 6.31) | < 0.001 | 3.40 (2.36, 4.41) | 4.24 (3.40, 6.83) | 0.002 |
| L (10<sup>9</sup>) | 1.12 (0.89, 1.54) | 0.66 (0.46, 0.95) | < 0.001 | 1.22 (0.91, 1.51) | 0.72 (0.50, 0.97) | < 0.001 |
| N/L | 2.69 (1.72, 4.45) | 6.37 (4.08, 11.03) | < 0.001 | 2.72 (1.58, 4.51) | 6.53 (4.39, 11.54) | < 0.001 |
| CRP,<br>mg/dL | 0.85 (0.16, 2.84) | 6.10 (3.44, 10.15) | 0.001 | 0.74 (0.24, 3.01) | 4.77 (2.06, 6.58) | < 0.001 |
| ALT, U/L | 19.60 (13.90,<br>31.90) | 23.70 (14.80,<br>33.70) | 0.264 | 17.30 (13.30,<br>31.20) | 23.60 (14.28,<br>37.70) | 0.157 |
| AST, U/L | 20.00 (16.80,<br>28.30) | 31.60 (24.70,<br>43.00) | 0.061 | 19.00 (16.05,<br>27.40) | 27.95 (20.10,<br>50.05) | 0.056 |
| ALB, g/L | 39.74 (5.13) | 35.84 (4.86) | < 0.001 | 40.07 (1.78) | 35.22 (4.18) | < 0.001 |
| Cr, µmol/L | 59.30 (50.20,<br>73.40) | 72.10 (57.90,<br>92.10) | 0.012 | 64.80 (49.00,<br>75.25) | 73.20 (57.95,<br>86.30) | 0.036 |
| BUN,<br>mmol/L | 3.78 (3.06, 4.70) | 5.58 (4.10, 8.28) | 0.031 | 3.92 (3.01, 4.73) | 5.60 (3.94, 7.50) | < 0.001 |
| D-dimer,<br>µg/mL | 0.45 (0.22, 1.36) | 3.26 (0.83, 9.68) | < 0.001 | 0.56 (0.28, 1.19) | 3.45 (1.01, 9.25) | < 0.001 |

HBP: high blood pressure; T2DM: type 2 diabetes mellitus; CHD: coronary heart disease; COPD: chronic obstructive pulmonary disease; onset time: time from onset to admission (days); WBC: white blood cells; L: lymphocyte; N: neutrophils; ALT: alanine transaminase; AST: aspartate aminotransferase; ALB: serum albumin; Cr: serum creatinine; BUN: blood urea nitrogen; CRP: C-reactive protein

**Table 4.** Reclassification results of three sets

| Original | Disease level<br>(Non-severe, Severe) | Predicted group membership |  |
| --- | --- | --- | --- |
|  |  | Non-severe | Severe |
| Training set | Non-severe | 121 | 50 |
|  | Severe | 8 | 71 |
| Test sets | Non-severe | 60 | 29 |
|  | Severe | 4 | 34 |
| Full datasets | Non-severe | 181 | 79 |
|  | Severe | 12 | 105 |

Patients with N/L\*CRP\*D-dimer results of less than 5.32 were reclassified into the non-severe group and those with results of greater than or equal to 5.32 were reclassified into the severe group

**Table 5:**
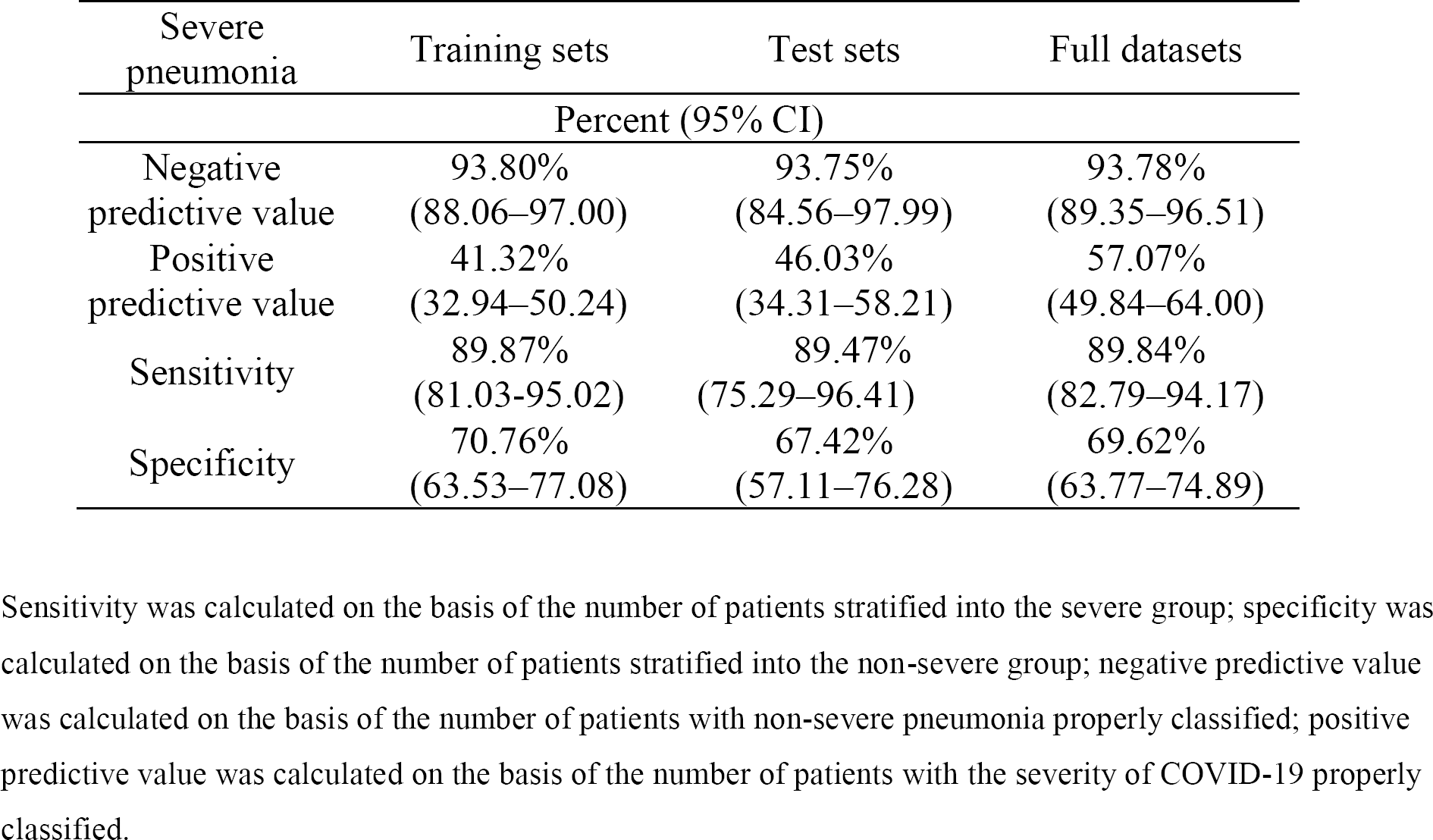
Results of internal validation

| Severe pneumonia | Training sets | Test sets | Full datasets |
| --- | --- | --- | --- |
| Percent (95% CI) |  |  |  |
| Negative predictive value | 93.80%<br>(88.06–97.00) | 93.75%<br>(84.56–97.99) | 93.78%<br>(89.35–96.51) |
| Positive predictive value | 41.32%<br>(32.94–50.24) | 46.03%<br>(34.31–58.21) | 57.07%<br>(49.84–64.00) |
| Sensitivity | 89.87%<br>(81.03–95.02) | 89.47%<br>(75.29–96.41) | 89.84%<br>(82.79–94.17) |
| Specificity | 70.76%<br>(63.53–77.08) | 67.42%<br>(57.11–76.28) | 69.62%<br>(63.77–74.89) |

Sensitivity was calculated on the basis of the number of patients stratified into the severe group; specificity was calculated on the basis of the number of patients stratified into the non-severe group; negative predictive value was calculated on the basis of the number of patients with non-severe pneumonia properly classified; positive predictive value was calculated on the basis of the number of patients with the severity of COVID-19 properly classified.

**Figure 5.**
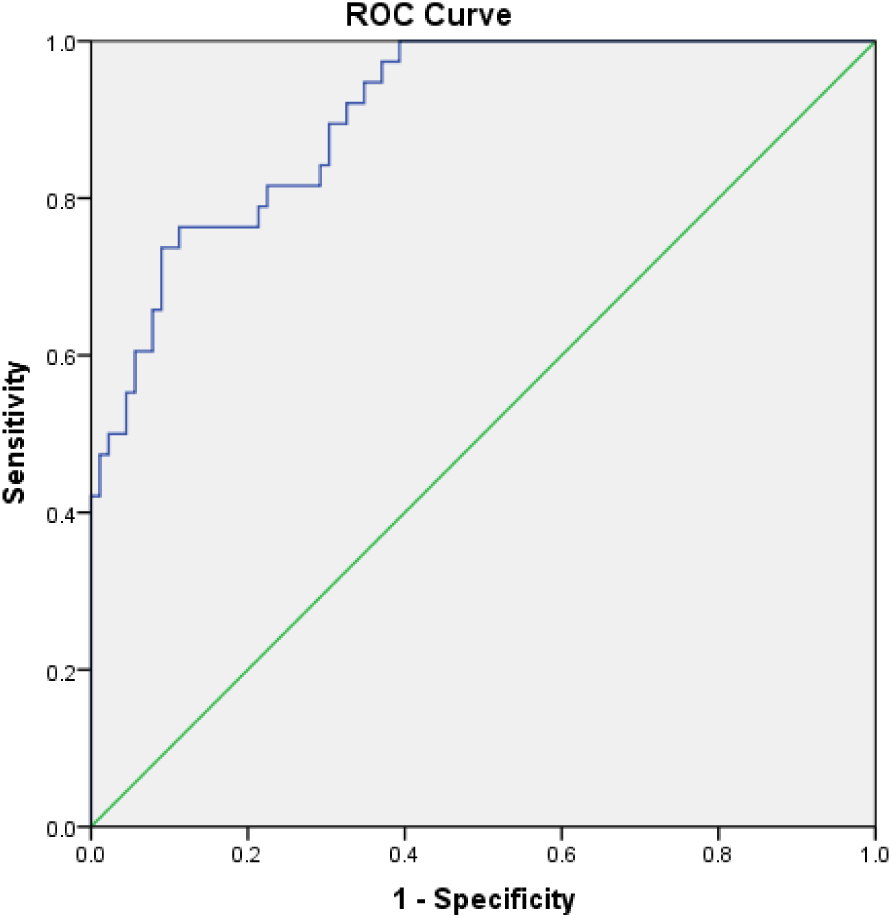
The receiver operating characteristic curve for N/L*CRP*D-dimer diagnosis of the severity of COVID-19 in the test sets; AUC: 0.906 (95% CI: 0.855–0.957).

**Figure 6.**
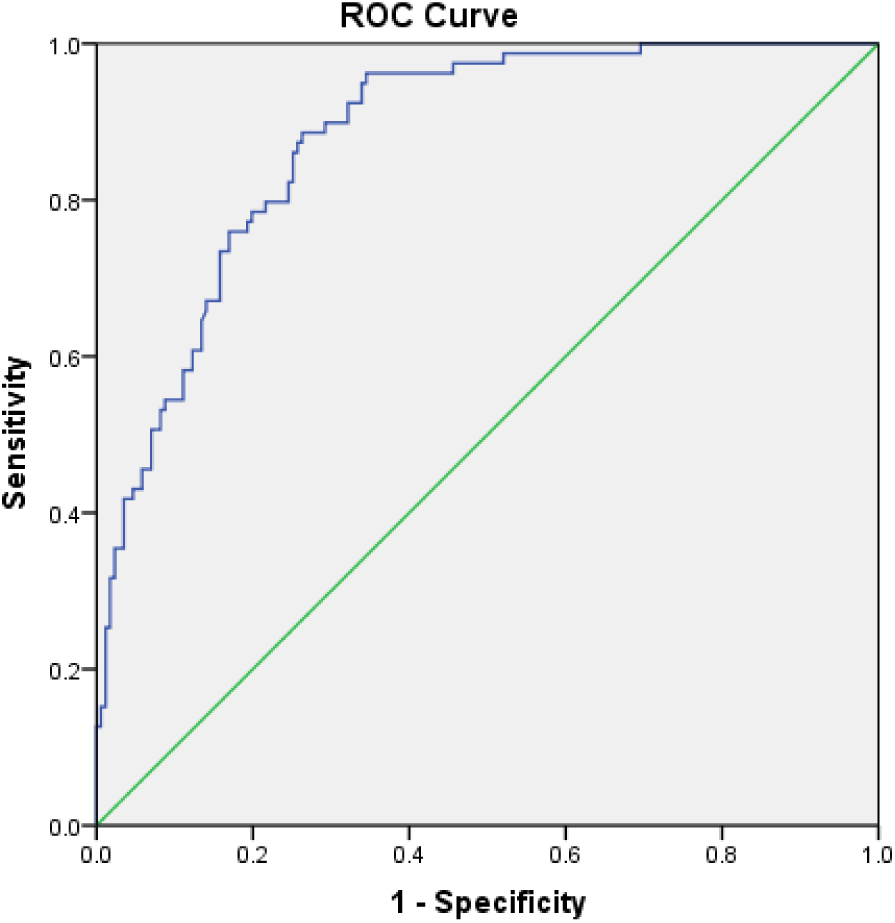
The receiver operating characteristic curve for N/L*CRP*D-dimer diagnosis of the severity of COVID-19 in the training sets; AUC: 0.879 (95% CI: 0.838–0.921).

In the training sets, the negative and positive predictive values were 93.80% (95% CI: 0.8806–0.9700) and 41.32% (95% CI: 0.3294–0.5024), respectively, with a specificity of 70.76% (95% CI: 0.6353–0.7708) and a sensitivity of 89.87% (95% CI: 0.8103–0.9502) (Tables 4 and 5). Results for negative and positive predictive values with the use of the full data set (training and test sets) are also shown in Tables 4 and 5.

## Discussion

This was a cross-sectional study that sought to investigate predictive factors of the severity of COVID-19. We found that age, N/L, CRP, and D-dimer were independent risk factors for the severity of COVID-19. Our results suggested that age was positively correlated with the severity of COVID-19. The result was also confirmed in other research^13^. This is probably because the resistance of the elders against virus decreases with age increase, basic diseases, and many other factors. Moreover, the aggravation of the basic disease profile of elderly patients would also lead to deterioration in the face of the severe pneumonia. The increases in N/L and CRP may be related to cytokine storms, which produce a series of inflammatory responses and cause disorders in peripheral WBCs and immune cells such as lymphocytes^14^. The observed decrease in lymphocytes indicated that coronavirus consumes many immune cells and inhibits the body’s cellular immune function. Damage to lymphocytes might contribute to exacerbations of patients and even cause death^15^. A significant increase of D-dimer in the progression of severe pneumonia was also observed, which was consistent with the results of Wang^13^. This may be correlated with the damage to the pulmonary arteries caused by the virus^16^, which leads to significant embolization in the extensive alveolar terminal capillaries. These changes eventually contribute to an increase in D-dimer.

Although N/L, CRP, and D-dimer were independent risk factors for the severity of COVID-19, the ROC curve showed that they have a low predictive value for the severity of the infection. In clinical practice, a very high negative predictive value is crucial in the evaluation of a patient with the severity of COVID-19, since failure to detect severe patients could have devastating consequences for these individuals. This study suggested that the product of N/L*CRP*D-dimer had better predictive performance for the severity of COVID-19 relative to single biomarkers. The predictive value of N/L*CRP*D-dimer was also confirmed in the internal validation stage. The observed negative predictive value of N/L*CRP*D-dimer was more than 90%, while the positive predictive value was more than 40%, with a specificity of 69.62% and a sensitivity of 89.84%, which appears to represent an improvement in prediction as compared with other clinical variables.

In general, the product of N/L*CRP*D-dimer is a new predictive value for the severity of COVID-19. The study validated an N/L*CRP*D-dimer cutoff point of 5.32 for the prediction. Clinicians should pay close attention to these indicators and calculate the value to identify severe patients as early as possible.

Importantly, our study has limitations. First, laboratory testing methods may not be the same in every hospital and the optimal cutoff point for the product value may vary. Further, only 377 patients were included in this study, where the product of N/L*CRP*D-dimer was found for the first time, so larger-scale data from randomized trials are needed to estimate whether to use this predictor in clinical practice.

## Data Availability

All data during the study are available in a repository or online.

## Acknowledgments

We thank LetPub (www.letpub.com) for its linguistic assistance during the preparation of this manuscript. Thanks to Mo lei from the medical science department of Shanghai lejiu medical technology co., LTD., for your help on statistics.

## Author contributions

Y.Z., Z.Y. and Y.N.G. had full access to all of the data in the study and take responsibility for the integrity of the data and the accuracy of the data analysis. Study concept and design: Y.F.W., Y.H. Acquisition, analysis, or interpretation of data: Y.F.W., Y.Z., Z.Y., Y.N.G., S.L.Y.,S.G. Drafting of the manuscript: Y.F.W., Y.Z., Z.Y. Critical revision of the manuscript for important intellectual content: all authors. Statistical analysis: Y.F.W, Y.Z. Administrative, technical, or material support: S.G., Z.Y. Study supervision: Y.F.W.and Y.H.

No author had any financial/non-financial disclosure.

No sponsor had any role.

## Abbreviation List

ALB: serum albumin
ALT: alanine transaminase
ARDS: acute respiratory distress syndrome
AST: aspartate aminotransferase
AUC: area under the receiver operating characteristic curve
BUN: blood urea nitrogen
COVID-19: coronavirus disease-19
Cr: serum creatinine
CHD: coronary heart disease
CRP: C-reactive protein
CT: computed tomography
HBP: high blood pressure
L: lymphocyte
N: neutrophil
N/L: ratio of neutrophils to lymphocytes
N/L*CRP*D-dimer: product of N/L, CRP, and D-dimer
ROC: receiver operating characteristic
SARS: severe acute respiratory syndrome
SARS-CoV-2: severe acute respiratory syndrome (SARS) coronavirus 2
T2DM: type 2 diabetes mellitus
WBC: white blood cell

